# Development & Deployment of a Real-time Healthcare Predictive Analytics Platform

**DOI:** 10.1101/2023.04.10.23288373

**Authors:** Aaron Boussina, Supreeth Shashikumar, Fatemeh Amrollahi, Hayden Pour, Michael Hogarth, Shamim Nemati

## Abstract

The deployment of predictive analytic algorithms that can safely and seamlessly integrate into existing healthcare workflows remains a significant challenge. Here, we present a scalable, cloud-based, fault-tolerant platform that is capable of extracting and processing electronic health record (EHR) data for any patient at any time following admission and transferring results back into the EHR. This platform has been successfully deployed within the UC San Diego Health system and utilizes interoperable data standards to enable portability.

**Clinical relevance:** This platform is currently hosting a deep learning model for the early prediction of sepsis that is operational in two emergency departments.

## I. INTRODUCTION

Despite the rapid growth in the number of predictive models developed for healthcare applications, there has been a relative dearth of successful implementations into clinical practice [1, 2]. One major reason for this is the large technical barrier to accessing EHR data in real-time and providing timely results back to clinicians [3, 4]. The challenges are multifactorial and include security, interoperability, availability, and scalability. Existing clinical decision support (CDS) solutions tend to be EHR-vendor or hospital specific and have not been generalized to new institutions [5]. Further, customized on-premise solutions to EHR integration are susceptible to system interruptions and are difficult to scale. In this work, we present a secure, high-availability, cloud-based platform that can process EHR data on any patient within a hospital at any point during their admission. We demonstrate how this platform can close the CDS loop and provide realtime recommendations to clinicians natively within the EHR. We further describe how process control tooling is leveraged to ensure system availability and model fidelity. Finally, we showcase the deployment of a deep learning model for the early prediction of sepsis onto this platform and into clinical practice [6].

The deployment of this sepsis model is clinically significant since sepsis (a life-threatening condition arising from the body’s overwhelming response to infection) is a major cause of mortality and morbidity globally [7-10]. The early recognition and treatment of sepsis has been shown to significantly improve outcomes [11-13]. The use of deep learning at the patient bedside can assist with risk stratification for sepsis management and has the potential to improve clinical outcomes.

This work is distinct from prior publications [14-15] in real-time healthcare analytics in the following ways: (1) It describes a platform for real-time predictions across all inpatient settings including Intensive Care Units (ICUs), Emergency Departments (EDs), and wards. (2) It utilizes collection of data elements beyond laboratory results, vital-signs, and demographics information, including problem-list items, procedures, and clinical notes. (3) It closes the CDS loop and enables outputs from predictive models to be written directly to the EHR. (4) It demonstrates the deployment of the platform into clinical practice.

## II. METHODS

### A. Platform Architecture

Figure 1 shows the high-level architecture for the predictive analytics platform. Data are extracted from the EHR at routine intervals and subsequently preprocessed for model consumption. An Indications-for-Use module assesses the clinical context of the patient including where the patient is in their treatment timeline to determine whether the patient is included/excluded for CDS. The predictive model is then run and outputs are sent back to the EHR to provide clinicians with relevant recommendations. The system is built in a modular plug-and-play manner such that any number of predictive modules can be added.

**Fig. 1.**
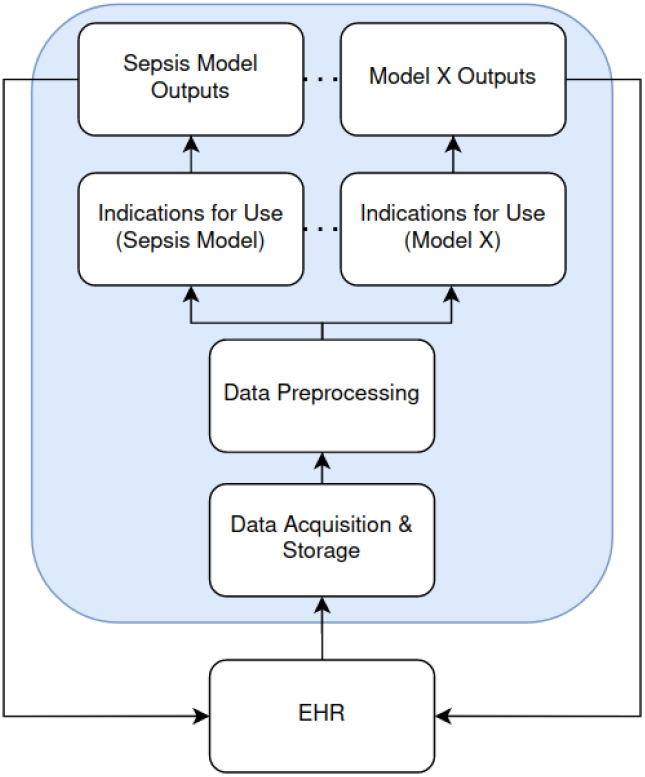
Architecture of the predictive analytics platform.

### B. Cloud Implementation

Our predictive analytics platform is hosted within a HIPAA-compliant Amazon Web Services (AWS) environment (Figure 2). The environment is an isolated enclave with communications only permitted through whitelisted ports. The application layer is hosted on a single EC2 instance with data stored in a MySQL Relational Database Service (RDS). The EC2 instance is part of an Auto Scaling Group (ASG) that is connected to a Network Load Balancer (NLB). This configuration enables a new copy of the EC2 instance to be brought online immediately in the event of primary instance failure. Similarly, the RDS is configured for regular backup which enables switchover to the secondary database in the event of failure.

**Fig. 2.**
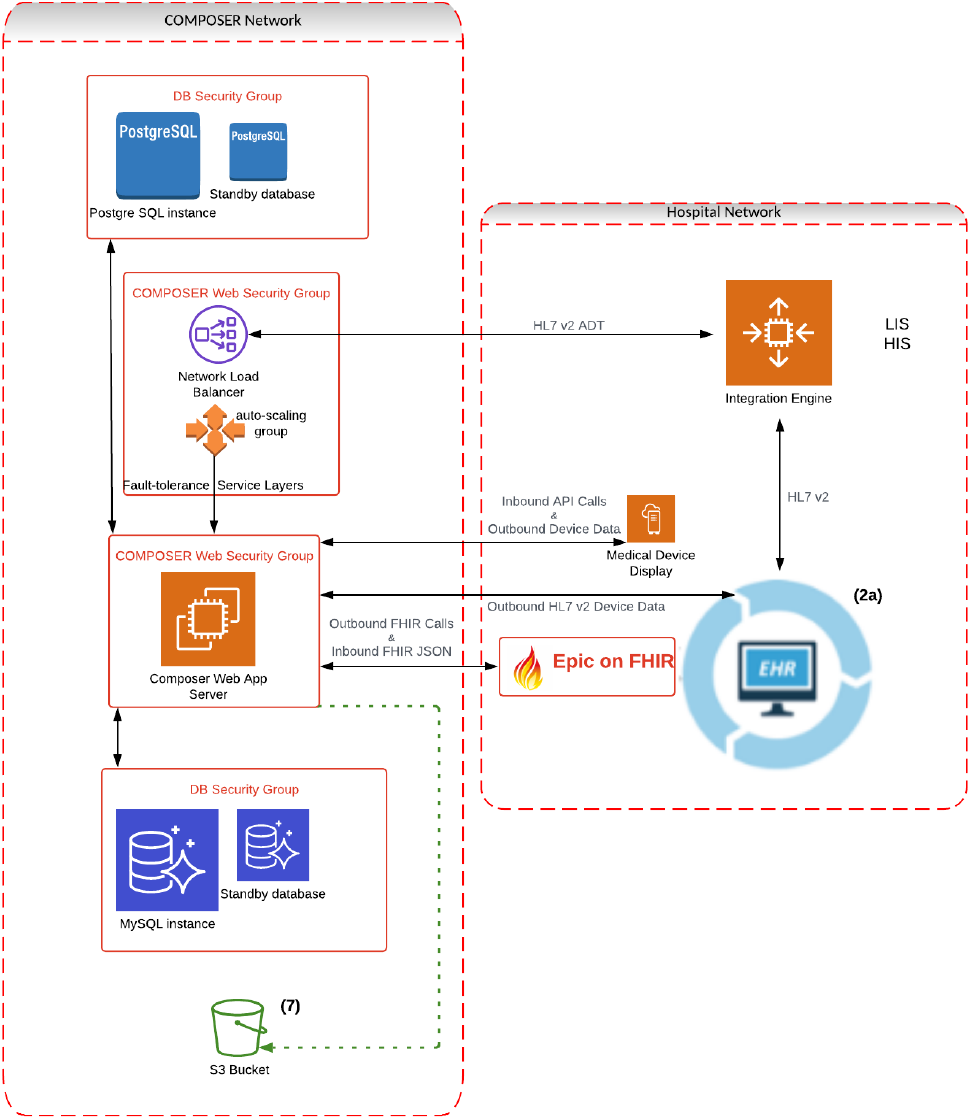
AWS implementation of the analytics platform.

For security, all system credentials such as private keys are stored within the AWS Secrets Manager and automatically updated on a routine schedule. For traceability, all system logs are preserved within S3 storage buckets. For portability, the architecture is captured within terraform scripts that automate the AWS build.

### C. Data Pipeline

All active admitted patients within the healthcare system are identified from HL7v2 ADT messages [16]. All ADT messages are forwarded from the hospital’s integration engine to MirthConnect software running within the application EC2 instance. The patient contact serial numbers (CSNs) from the PID segment are converted to patient FHIR IDs by calling the “Patient.Search” API from the hospital’s Epic FHIR server. The platform application authenticates its requests to the FHIR server with OAuth 2.0 using a backend private key.

With the patient FHIR IDs retrieved for every admitted patient, the application makes regular calls to the FHIR server to retrieve updates to the Patient, ServiceRequest, Observation, MedicationRequest, Condition, and Procedure resources. The resources are returned as JSON bundles which are then parsed and preserved within the RDS as a condensed JSONB column containing all updated data for a patient within an elapsed timeframe.

The semi-structured data are then passed to the Data Preprocessing Module where they are converted into a structured format with columns for every feature and missing values imputed by a predefined sample-and-hold. The Data Preprocessing Module also enforces that the values are physiologically possible and not the result of inadvertent entry by applying upper and lower limits on the features.

Models are then directly deployed using these database tables as inputs. Model outputs such as the risk score and top contributing features are placed into OBX segments and an outbound HL7v2 message is constructed and sent back to the integration engine. The observation identifier (field OBX.3) is registered within the EHR allowing all model outputs to be filed to the flowsheet. EHR-native decision support, such as Best Practice Advisories (BPAs), then utilize these flowsheet items to generate clinician-facing alerts.

### D. Model Deployment

Using this platform, we deployed the COMPOSER deep learning model for the early prediction of sepsis described in [6]. The model was run in silent-mode evaluation over the course of 6 months in which sepsis risk scores were filed to patient flowsheets, but alerts were not displayed to clinicians. During this period, the performance of the model was evaluated and routine chart reviews with a panel of experts were conducted to assess the clinical utility of the silent alerts. These reviews informed the development of the indications for use of this algorithm listed in Table 1.

**TABLE I.** Indications for Use of the COMPOSER Algorithm

|  |
| --- |
| Admitted to the ED |
| Age of $\geq 18$ |
| No code for hospice or comfort care |
| Not admitted for a planned procedure |
| Does not have 2 out of 3 of the following orders:<br>(1) Antibiotics, (2) Blood culture, (3) Lactate |
| No previous COMPOSER alerts for the patient |

Design sessions with nursing teams were conducted over the span of three months to build the display of the final EHR-native BPA (Figure 3). Following prospective validation of the model performance, training was conducted across two emergency departments within the UC San Diego Health system prior to deployment of the BPA into clinical workflow.

**Fig. 3.**
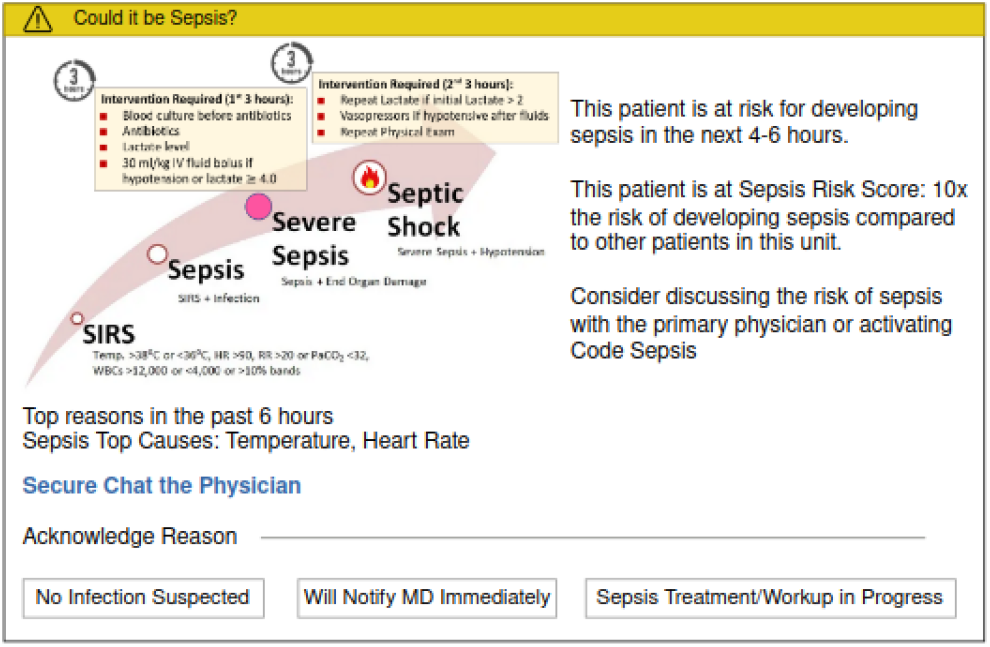
Sample display of the COMPOSER BPA.

### E. Process Control

To ensure high-availability of the platform, AWS CloudWatch alerts were created to notify the development team of any event that brought the system out of a state of control (e.g. if a service was unreachable). These CloudWatch alerts were integrated with PagerDuty to ensure 24/7 front-line support. Further, the platform was registered with healthcare IT within ServiceNow to enable end-users to escalate issues directly to the development team.

In addition to system interruptions, deployed models are at risk of model drift due to changes in the data distribution over time [17-19]. To ensure detection of possible model drift we implemented a quality dashboard that automatically tracks a model’s inputs, outputs, and performance. Specifically, the median values of measurement inputs and risk score outputs are monitored to ensure they don’t pass the upper or lower quartiles from the training cohort. If any value falls outside of those limits, it is flagged for review by the development team. Similarly, model performance metrics such as the positive predictive value (PPV) and sensitivity are tracked on a weekly basis. Finally, the quality dashboard tracks the rate of rejection from conformal prediction [6]. Conformal prediction is a method for detecting out-of-distribution data within the lower dimensional representations of a neural network. Changes in rejection rates, therefore, are expected to correspond to changes in data distribution that affect the model’s predictions.

## III. RESULTS

### A. Clinical Population

Table 2 shows the patient population processed by the platform from June 1st, 2022 to January 1st, 2023. During this 7-month period, 63,133 patients and 1,368,763 patient-hours were processed across 63 care units.

**TABLE II.** Demographics and clinical characteristics of patients processed by the real-time analytics platform.

|  |  |
| --- | --- |
| Number of Patients, N | 63,133 |
| Age, Mean (SD) | 52.0 (19.08) |
| Males, N (%) | 31,187 (49.4%) |
| Females, N (%) | 31,860 (50.5%) |
| Unknown Gender, N (%) | 86 (0.14%) |
| ED Patients, N | 47,993 |
| ICU Patients, N | 3,565 |
| Ward Patients, N | 14,185 |
| Length of Stay (hours), Median (IQR) | 6.7 [4.03 - 8.47] |

### B. Sepsis Model Performance

Figure 4 shows outputs exported from the quality dashboard for the median values of a sample input feature and COMPOSER’s sepsis risk score relative to the training set following clinical deployment on 2022-12-07. Also shown are the model’s PPV and rate of conformal rejection. The dashboard demonstrates that the input feature distributions and model outputs had not drifted substantially post-deployment. Further, the model’s performance in real-time using the analytics platform did not differ significantly from retrospective performance.

**Fig. 4.**
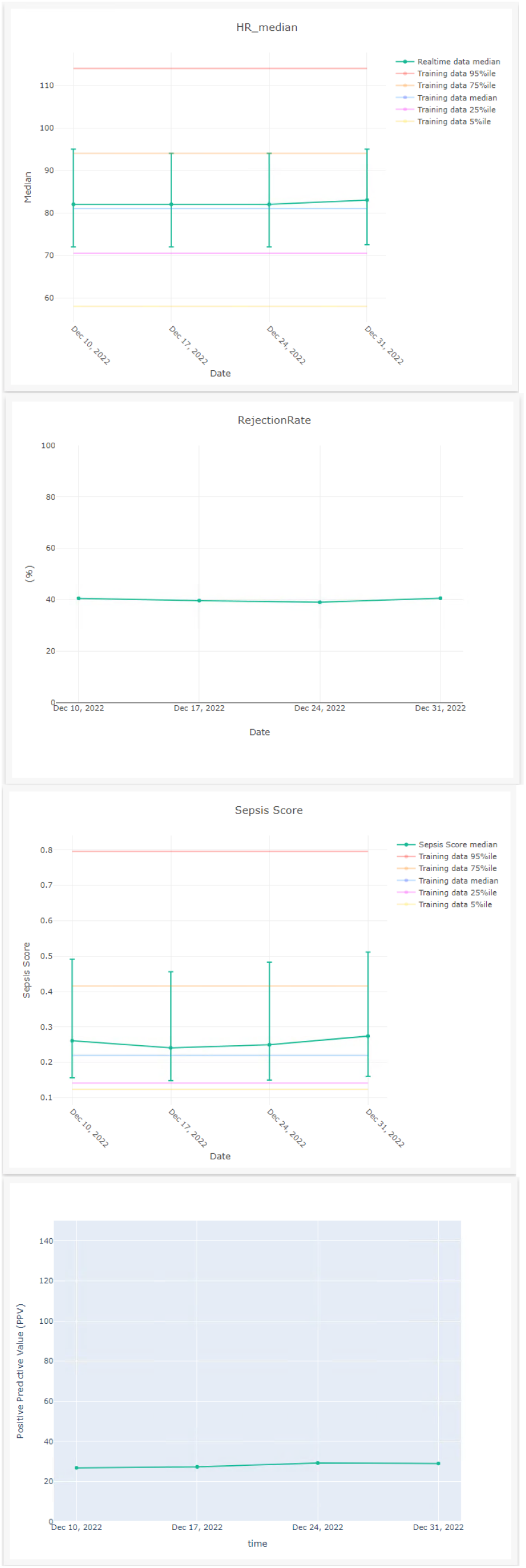
Median values and 95% confidence intervals of input heart rate feature and output COMPOSER risk score relative to the training dataset. Trends of conformal rejection rate and PPV over time.

### C. System Availability

From June 1st, 2022 to January 1st, 2023 the platform experienced a total of 0 hours of system downtime and 28 hours of inter-connectivity interruptions. This corresponds to an overall uptime of 99.44%. The single largest instance of downtime (10 hours) was related to a significant update to the FHIR API protocol which resulted in a difference in version parity with the platform.

## IV. CONCLUSIONS

Using cloud architecture and interoperable data standards we have built a production-grade system to enable safe, rapid deployment of predictive analytics into the clinic. We have leveraged best-practices in software engineering and process control to ensure that the platform is sustainable and robust. We have designed the platform together with our clinical collaborators to ensure that model predictions are relevant and clinically actionable. This work aims to address the growing divide between the abundance of new deep learning models and the relative paucity of predictive models in clinical practice. While in this work we have only showcased a single model at a single institution, we have developed the system with portability and scalability in mind. We are currently developing prediction models for other clinical use cases on this platform and targeting additional institutions for deployment.

## Data Availability

All data produced in the present work are contained in the manuscript.

## ACKNOWLEDGMENT

S.N. is funded by the National Institutes of Health (#R01LM013998, #R01HL157985, #R35GM143121). He is co-founder of a UCSD start-up, Healcisio Inc., which is focused on commercialization of advanced analytical decision support tools. Mr. Boussina is funded by the National Library of Medicine (#2T15LM011271-11). Dr. Shashikumar has no sources of funding to declare. The opinions or assertions contained herein are the private ones of the author and are not to be construed as official or reflecting the views of the NIH or any other agency of the US Government.

